# Electronic health record biobank cohort recapitulates an association between the *MUC5B* promoter polymorphism and ARDS in critically ill adults

**DOI:** 10.1101/2024.08.26.24312498

**Authors:** V. Eric Kerchberger, J. Brennan McNeil, Neil Zheng, Diana Chang, Carrie Rosenberger, Angela J. Rogers, Julie A. Bastarache, QiPing Feng, Wei-Qi Wei, Lorraine B. Ware

## Abstract

**Background:** Large population-based DNA biobanks linked to electronic health records (EHRs) may provide novel opportunities to identify genetic drivers of ARDS.

**Research Question:** Can we develop an EHR-based algorithm to identify ARDS in a biobank database, and can this validate a previously reported ARDS genetic risk factor?

**Study Design and Methods:** We analyzed two parallel genotyped cohorts: a prospective biomarker cohort of critically ill adults (VALID), and a retrospective cohort of hospitalized participants enrolled in a de-identified EHR biobank (BioVU). ARDS was identified by clinician-investigator review in VALID and an EHR algorithm in BioVU (EHR-ARDS). We tested the association between the *MUC5B* promoter polymorphism rs35705950 with development of ARDS, and assessed if age modified this genetic association in each cohort.

**Results:** In VALID, 2,795 patients were included, age was 55 [43, 66] (median [IQR]) years, and 718 (25.7%) developed ARDS. In BioVU, 9,025 hospitalized participants were included, age was 60 [48, 70] years, and 1,056 (11.7%) developed EHR-ARDS. We observed a significant age-related interaction effect on ARDS in VALID: among older patients, rs35705950 was associated with increased ARDS risk (OR: 1.44; 95%CI 1.08-1.92; p=0.012) whereas among younger patients this effect was absent (OR: 0.84; 95%CI: 0.62-1.14; p=0.26). In BioVU, rs35705950 was associated with increased risk for EHR-ARDS among all participants (OR: 1.20; 95%CI: 1.00-1.43, p=0.043) and this did not vary by age. The polymorphism was also associated worse oxygenation in mechanically ventilated BioVU participants, but had no association with oxygenation in VALID.

**Interpretation:** The *MUC5B* promoter polymorphism was associated with ARDS in two cohorts of at-risk adults. Although age-related effect modification was observed only in VALID, BioVU identified a consistent association between *MUC5B* and ARDS risk regardless of age, and a novel association with oxygenation impairment. Our study highlights the potential for EHR biobanks to enable precision-medicine ARDS studies.

## INTRODUCTION

The acute respiratory distress syndrome (ARDS) is an acute inflammatory lung disorder that develops in the setting of a local or systemic insult such as pneumonia, sepsis, or massive trauma.^1^ ARDS occurs in over 20% of mechanically ventilated critically ill adults, and leads to prolonged hospitalization, high costs, and high mortality risk. Understanding genetic drivers of ARDS among at-risk individuals could improve our ability to define disease subphenotypes, predict disease risk, and identify new therapeutic targets.^2^

As the risk for ARDS is influenced by many genes, each exerting only a modest contribution to an individual’s overall risk, large numbers of cases and at-risk control patients are necessary to design appropriately powered studies.^2–6^ However, prospectively identifying and genotyping thousands of critically ill patients and phenotyping them specifically for ARDS is prohibitively costly and time consuming. Therefore, innovative sampling approaches are needed to design adequately powered genetic studies in the ICU. Large population-based DNA biobanks pairing clinical data from the electronic health record (EHR) with genomic information have improved our understanding of genetic contributors to many chronic diseases.^7^ However, leveraging EHR biobanks to study ARDS requires novel methods because of its syndromic definition.^8–11^

We previously reported that a gain-of-function polymorphism (rs35705950) in the gel-forming mucin gene *MUC5B* promoter was associated with increased risk for ARDS among critically ill adults aged 50 years or older.^12^ The original study included 1,534 adults (including 903 aged ≥50 years) enrolled from a prospective observational ICU cohort and genotyped using a dedicated polymerase-chain reaction (PCR) assay. In this study, we have expanded genotyping of this prospective ICU cohort to nearly 3,000 patients on a single genome-wide array platform (which includes patients in the original study), increasing our power to detect genetic associations with ARDS. Then to validate the association between rs35705950 and ARDS, we developed a parallel cohort from our institution’s de-identified EHR biobank by employing a rule-based ARDS classifier based on a combination of diagnostic billing codes, laboratory test results, and text from unstructured radiography reports. We analyzed both cohorts in a derivation-validation approach to test the association between ARDS risk and rs35705950, and hypothesized that the *MUC5B* polymorphism would exert differential risk for developing ARDS based on age.

## MATERIALS AND METHODS

### Patient Cohorts

A study schematic is provided in **Figure 1**. The derivation cohort came from the Validating Acute Lung Injury biomarkers for Diagnosis (VALID) study, a prospective observational cohort of critically ill adults at risk for ARDS, admitted to either the surgical, medical, trauma, or cardiovascular intensive care units (ICUs) at Vanderbilt University Medical Center (VUMC) from January 2006 to December 2020. All patients were enrolled on the morning of the second ICU day. Other inclusion criteria, enrollment and consent procedures for VALID have been previously described.^9,13^ Two expert physician investigators manually reviewed clinical records and chest radiographs to adjudicate ARDS status according to the Berlin Definition,^14^ and we defined ARDS cases as those with expert-adjudicated ARDS on any of the first four ICU days.

The validation cohort was identified from BioVU, VUMC’s DNA-biobanking program linked to a de-identified EHR (**Figure 1**).^7,15^ Additional information on the BioVU enrollment process and the de-identified EHR are provided in the **Supplementary Methods**. We identified BioVU participants age ≥18 years admitted to the hospital from 2002 to 2019 with at least one diagnostic billing code for an ARDS risk factor during the first 7 days of hospital admission (**e-Table 1**). We included both ICU and non-ICU participants as many patients with ARDS risk factors who ultimately do not develop the syndrome are managed outside of the ICU setting. We then developed a computable ARDS algorithm (EHR-ARDS) to capture the four criteria of the Berlin ARDS Definition using structured EHR data.^14^ EHR-ARDS cases were identified by presence of (i) a diagnostic billing code for acute hypoxemic respiratory failure (**e-Table 2**), (ii) procedural codes for mechanical ventilation (**e-Table 3**), (iii) PaO2:FiO2 ratio ≤300, (iv) presence of a chest x-ray report containing terms consistent with bilateral opacities as identified using a set of regular expressions that we designed *a priori* for the study (**e-Table 4**), and (v) more days with diagnosis codes for ARDS risk factors than days with codes for congestive heart failure (**e-Table 5**). At-risk controls included all participants who were admitted with an ARDS risk factor diagnosis code but did not meet all EHR-ARDS criteria during the first 7 days of hospitalization. For participants with more than one qualifying admission, we used the first hospitalization only. Patients/participants with missing genotypes or diagnosis codes for interstitial lung diseases (ILDs), lung transplant, or bone marrow transplant were excluded from both cohorts (**e-Table 6**). The Vanderbilt Institutional Review Board (Nashville, TN) reviewed and approved the study protocols for both the VALID (IRB #051065) and BioVU cohorts (IRB # 202530).

**Figure 1:**
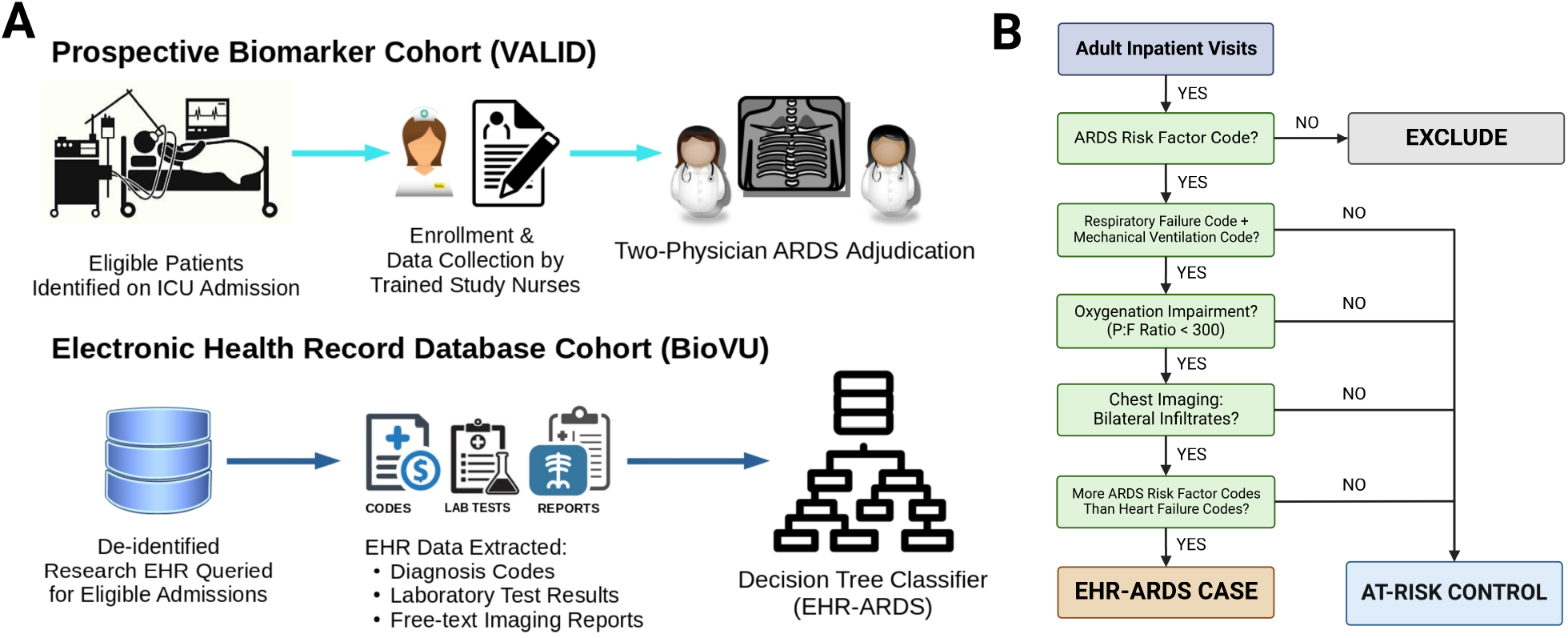
Schematic of Study Cohorts and EHR-ARDS Classifier. Schematic diagrams of (A) Study Cohorts and (B) EHR-ARDS Classifier.

### EHR-ARDS Classification Performance

We assessed the EHR-ARDS classifier’s performance in both cohorts using standard metrics including sensitivity, specificity, positive predictive value (PPV), negative predictive value (NPV), F_1_ score, and Cohen’s κ-statistic.^16^ In VALID, we assessed performance of EHR-ARDS against investigator-adjudicated Berlin ARDS among all genotyped patients included in the study. In the BioVU cohort, EHR-ARDS classifier performance was compared to ARDS status for 125 randomly selected participants (50 EHR-ARDS, 75 at-risk control participants) by manual adjudication of Berlin ARDS status using clinical notes and radiography reports available in the de-identified EHR’s user interface.^15^

### *MUC5B* Promoter Genotyping

VALID patients were genotyped using the Illumina Global Screening Array and BioVU participants were genotyped using the Illumina Expanded Multi-Ethnic Global Array. We ascertained *MUC5B* promoter (rs35705950) genotype in both cohorts by imputation using the Michigan Imputation Server with the Haplotype Reference Consortium version 1.1 reference panel.^17,18^ Additional genotyping information is provided in the **Supplementary Methods**.

### Statistical Analysis

Continuous variables are presented as median and interquartile ranges, categorical variables as frequencies and proportions, and differences between groups as mean differences and 95% confidence intervals (CI). We assessed group-wise differences using the Mann-Whitney *U* test for continuous outcomes and Pearson’s χ^2^-test for categorical outcomes. We tested the association between the *MUC5B* promoter polymorphism and outcomes using logistic regression for ARDS in VALID or EHR-ARDS in BioVU, linear regression for lowest PaO_2_:FiO_2_ ratio, and ordinal regression for Berlin ARDS severity categories.^14^ In-hospital survival was assessed using the method of Kaplan and Meier. As we used imputed genotypes for rs35705950, genotype was treated as a continuous variable ranging from 0.0 to 2.0 to represent the predicted number of alterative (T) alleles and to account for uncertainty in the imputed gene dosage. We selected model covariates using a causal directed acyclic graph (**e-Figure 1**),^19^ which included age, sex, race, ARDS risk factors (aspiration, pancreatitis, pneumonia, sepsis, shock, or trauma), and cardiovascular comorbidities (heart failure, acute myocardial infarction, or other coronary artery disease). Based on the age-dependent effect of the promoter polymorphism observed among VALID patients in the previous study,^12^ we assessed the associations between genotype and ARDS/EHR-ARDS both without any interaction terms and with including a genotype × age interaction term in the regression model. The combined significance of the *MUC5B* genotype and the genotype × age interaction term was also assessed using an F-test on both parameters. A p-value of less than 0.05 was considered statistically significant as all analyses were considered exploratory. All regression models employed heteroscedasticity-consistent standard errors using the “HC3” covariance matrix estimators.^20,21^ We performed all analyses using R version 4.3.2 (Vienna, Austria) with the R packages *interactions* and *sandwich*.^21,22^

## RESULTS

### Study Populations

Demographic and clinical characteristics of both cohorts are shown in **Table 1** and flow diagrams illustrating patients meeting inclusion and exclusion criteria are shown in **e-Figure 2**. Compared to VALID, the BioVU cohort was older (median [IQR] age: 60 [48, 70] years vs 55 [43, 66] years, p<0.001), had a more even sex distribution (male sex: 49.2% vs 60.1%, p<0.001), had lower rates of most ARDS risk factor diagnoses, and had lower rates of mechanical ventilation (20.7% vs 76.1%, p<0.001). Hospital length of stay and inpatient mortality were also lower in the BioVU cohort, reflecting inclusion of participants hospitalized in both ICU and non-ICU settings. *MUC5B* promoter polymorphism T-allele frequencies were similar between cohorts (VALID: 10.5% BioVU: 9.6%, p-value: 0.15, **e-Figure 3**). We observed high agreement in VALID between promoter genotype determined by imputation and by polymerase chain reaction (**e-Table 7**, Cohen’s κ: 0.98).

**Table 1:** Study Population Demographics.

| Variable | Prospective cohort (VALID) | EHR Cohort (BioVU) |
| --- | --- | --- |
| **Demographics** |  |  |
| Number | 2,795 | 9,025 |
| Age (years) (median [IQR]) | 55 [43, 66] | 60 [48, 70] |
| Age over 50 years (%) | 1,753 (62.7) | 6,413 (71.1) |
| Sex (%) |  |  |
| Male | 1,679 (60.1) | 4,442 (49.2) |
| Female | 1,116 (39.9) | 4,583 (50.8) |
| Recorded Race (%) |  |  |
| White | 2,407 (86.1) | 7,602 (84.2) |
| Black | 350 (12.5) | 1,170 (13.0) |
| Asian | 5 ( 0.2) | 70 ( 0.8) |
| Multiracial | 0 ( 0.0) | 48 ( 0.5) |
| Other/Not Recorded | 33 ( 1.2) | 135 ( 1.5) |
| Recorded Ethnicity (%) |  |  |
| Non-Hispanic Non-Latino | 2,769 (99.1) | 8,754 (97.0) |
| Hispanic/Latino | 21 ( 0.7) | 157 ( 1.7) |
| Other/ Not Recorded | 5 ( 0.2) | 114 ( 1.3) |
| **Hospital Characteristics** |  |  |
| Admitted to ICU (%) | 2,795 (100.0) | 3950 (43.8) |
| Admitting ICU (%) |  |  |
| Burn | 0 (0.0) | 102 ( 2.6) |
| Cardiovascular | 80 ( 2.9) | 565 (14.3) |
| Medical | 1375 ( 49.2) | 1435 (36.3) |
| Neuroscience | 0 (0.0) | 346 ( 8.8) |
| Pediatrics | 0 (0.0) | 5 ( 0.1) |
| Surgical | 501 ( 17.9) | 1000 (25.3) |
| Trauma | 839 ( 30.0) | 497 (12.6) |
| **Respiratory Characteristics** |  |  |
| Mechanical Ventilation (%) | 2,128 (76.1) | 1,866 (20.7) |
| Worst PaO<sub>2</sub>/FiO<sub>2</sub> during observation (median [IQR]) | 161 [103, 240] | 137 [87, 195] |
| Any PaO<sub>2</sub>/FiO<sub>2</sub> Measured (%) | 2,110 (75.5) | 1,866 (20.7) |
| ARDS or EHR-ARDS (%) | 718 (25.7) | 1,056 (11.7) |
| Berlin ARDS Severity (%) <sup>a</sup> |  |  |
| Mild (PaO<sub>2</sub>/FiO<sub>2</sub> > 200 mmHg and ≤ 300 mmHg) | 78 (10.8) | 160 (15.1) |
| Moderate (PaO<sub>2</sub>/FiO<sub>2</sub> > 100 mmHg and ≤ 200 mmHg) | 363 (50.6) | 494 (46.8) |
| Severe (PaO<sub>2</sub>/FiO<sub>2</sub> ≤ 100 mmHg) | 277 (38.6) | 402 (38.1) |
| **Hospital Outcomes** |  |  |
| ICU Length of Stay (days) (median [IQR]) <sup>b</sup> | 6.0 [4.0, 11.0] | 4.0 [2.0, 9.0] |
| Hospital Length of Stay (days) (median [IQR]) | 12.0 [7.0, 20.0] | 5.0 [3.0, 9.0] |
| Inpatient Mortality (%) | 456 (16.3) | 537 ( 6.0) |
| **ARDS Risk Factors <sup>c</sup>** |  |  |
| Sepsis (%) | 1,312 (46.9) | 3,199 (35.4) |
| Pneumonia (%) | 1,242 (44.4) | 2,763 (30.6) |
| Trauma (%) | 987 (35.3) | 1,188 (13.2) |
| Shock (%) | 1,913 (68.4) | 4,656 (51.5) |
| Aspiration (%) | 363 (13.0) | 495 ( 5.5) |
| Pancreatitis (%) | 136 ( 4.9) | 749 ( 8.3) |
| **Cardiac Comorbidity** |  |  |
| Heart Failure (%) | 503 (18.0) | 1,509 (16.7) |
| Acute Myocardial Infarction (%) | 405 (14.5) | 1,151 (12.8) |
| Other Coronary Artery Disease (%) | 649 (23.2) | 2,201 (24.4) |
| **MUC5B Promoter Polymorphism Genotype (%)** |  |  |
| GG | 2,245 (80.3) | 7,388 (81.9) |
| GT | 513 (18.4) | 1,540 (17.1) |
| TT | 37 ( 1.3) | 97 ( 1.1) |

<sup>a</sup> Proportion of ARDS patients or EHR-ARDS participants
<sup>b</sup> Among patients/participants admitted to an ICU
<sup>c</sup> Patients/participants could have more than one ARDS risk factor

### EHR-ARDS Classifier Performance

Performance of the EHR-ARDS classifier was acceptable. In VALID, the classifier had a sensitivity 0.86, specificity 0.70, PPV 0.49, NPV 0.93, and Cohen’s κ 0.45, indicating moderate agreement with investigator-adjudicated ARDS (**Table 2**). We observed similar performance compared with investigator-adjudicated ARDS among 125 randomly selected participants in the BioVU cohort, with an observed sensitivity 0.94, specificity 0.81, PPV 0.66, NPV 0.97, and Cohen’s κ 0.67 (**Table 2**). Among BioVU participants with EHR-ARDS, we observed a clear association with mortality based on Berlin severity criteria: mild EHR-ARDS participants had a 30-day in-hospital survival of 87% (95% CI: 0.82 to 0.93), while survival was 73% (68% to 80%) moderate EHR-ARDS and 66% (60% to 72%) for severe ARDS (**e-Figure 4**).

**Table 2:** Classification Performance of EHR-ARDS Classifier versus Clinician Review.

|  | EHR Classifier Predictions |  |  |  |
| --- | --- | --- | --- | --- |
|  | VALID |  | BioVU |  |
|  | EHR-ARDS | EHR At-Risk Control | EHR-ARDS | EHR At-Risk Control |
| ARDS by Clinician Review | 616 | 102 | 33 | 2 |
| Not ARDS by Clinician Review | 632 | 1,445 | 17 | 73 |
| Classification Metric <sup>a</sup> |  |  |  |  |
| Sensitivity | 0.86 (0.83-0.88) |  | 0.94 (0.81-0.98) |  |
| Specificity | 0.70 (0.68-0.72) |  | 0.81 (0.72-0.89) |  |
| Positive Predictive Value | 0.49 (0.47-0.52) |  | 0.66 (0.52-0.78) |  |
| Negative Predictive Value | 0.93 (0.92-0.95) |  | 0.97 (0.91-0.99) |  |
| F<sub>1</sub> Score | 0.63 (0.61-0.65) |  | 0.78 (0.71-0.85) |  |
| Cohen $\kappa$ | 0.45 (0.41–0.48) | | 0.66 (0.53-0.80) | |

<sup>a</sup> Presented as point estimates and (95% confidence intervals). Confidence intervals were calculated using the Wilson score method for Sensitivity, Specificity, Positive Predictive Value, Negative Predictive Value, and F1 Score; and using a Wald interval for the κ-statistic.<sup>16</sup>

### Elevated ARDS Risk among Older VALID Patients with *MUC5B* Promoter Polymorphism

Similar to our previous report from VALID,^12^ we found no significant association between rs35705950 and ARDS in the overall cohort when not accounting for any age-related effect modification (OR per T allele: 1.11; 95% CI: 0.90-1.36; p-value: 0.35). In contrast, under the interaction analysis we observed a significant age × genotype interaction effect such that the effect of rs35705950 on ARDS increased among older patients (OR for interaction term [per 10 years increased age]: 1.17, 95% CI: 1.04-1.33, p-value: 0.015, joint rs35705950 and interaction term p-value: 0.026, **Figure 2**). For a patient aged 70 years (mean + 1 standard deviation for the cohort), each T-allele was associated with an odds ratio of 1.44 for ARDS (95% CI 1.08-1.92; p-value: 0.012), whereas for a patient age 37 years (mean - 1 standard deviation), the effect was substantially attenuated (OR per T-allele: 0.84; 95% CI: 0.62-1.14; p-value: 0.26). Subgroup analyses without the interaction term revealed more pronounced associations among older patients (age ≥ 50) compared to younger patients, females compared to males, and among patients who died during hospitalization compared to survivors, although the 95% confidence intervals did not exclude 1.0 for any analyzed subgroup (**e-Figure 5**).

### Elevated Risk of EHR-ARDS in BioVU Participants with *MUC5B* Promoter Polymorphism

In the BioVU cohort, we observed a significant association between rs35705950 and EHR-ARDS risk even without accounting for any interaction effects (OR per T-allele: 1.20; 95% CI: 1.00-1.43; p-value: 0.043). In contrast to VALID, the effect of the polymorphism on EHR-ARDS was consistent across all ages (OR for interaction term [per 10 years increased age]: 1.02, 95% CI: 0.91-1.13; p-value: 0.78; joint rs35705950 and interaction term p-value: 0.12, **Figure 2**).

**Figure 2:**
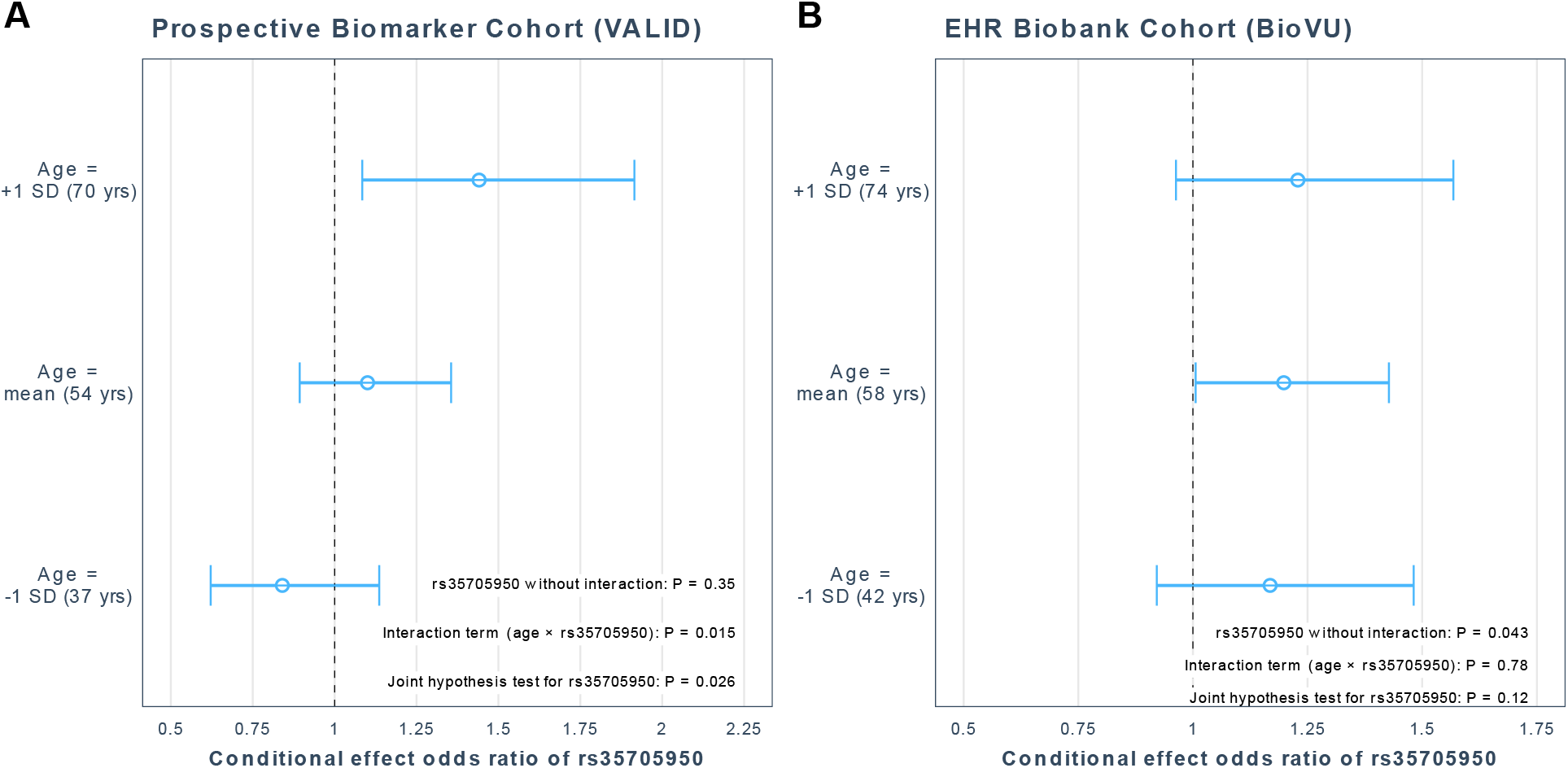
*MUC5B* promoter polymorphism is associated with increased ARDS risk. Conditional effects odds ratios for the *MUC5B* promoter polymorphism in (A) the prospective biomarker cohort (VALID), and (B) the EHR biobank cohort (BioVU). For each cohort, the y-axis indicates conditional odds ratios for (A) clinician-adjudicated ARDS or (B) EHR-ARDS per rs35705950 T-allele among younger (mean cohort age – 1 standard deviation), middle-aged (mean cohort age) and older (mean cohort age + 1 standard deviation) patients. Significant effect modification for age on rs35705950 was observed in VALID, but not in BioVU.

Subgroup analyses revealed that the association between rs35705950 and EHR-ARDS was generally consistent across all ARDS risk factor subgroups, although the association only reached our pre-specified level of statistical significance among the subgroup of participants hospitalized with shock (**Figure 3**). Similar to VALID, the association was also stronger among older patients (age ≥ 50) compared to younger patients, females compared to males, and among patients who died during the hospitalization compared to survivors (**Figure 3**).

**Figure 3:**
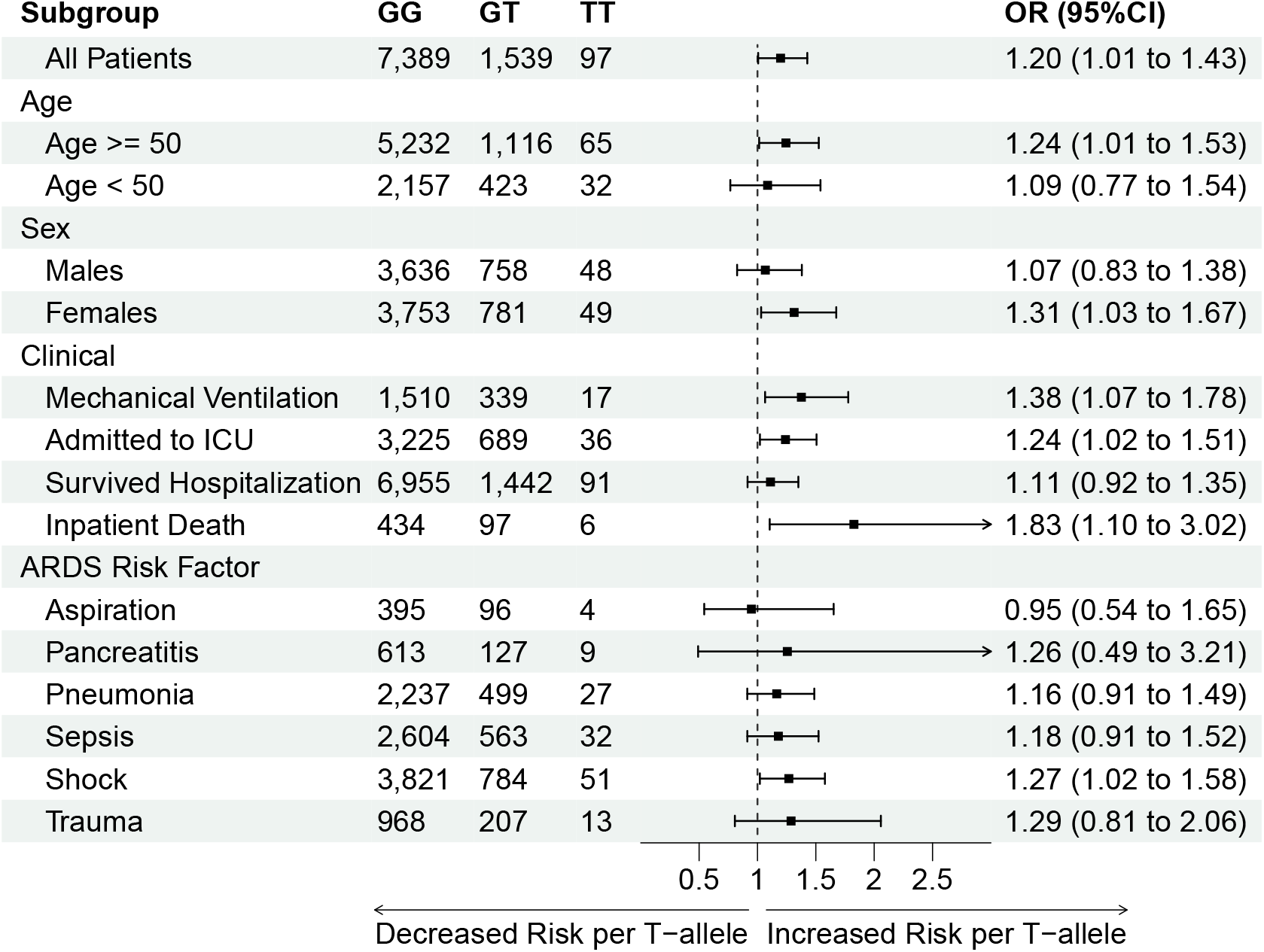
Subgroup Analysis of *MUC5B* Promoter Polymorphism and EHR-ARDS Risk in BioVU Cohort. Decreased Risk per T-allele Increased Risk per T-allele The odds ratio and 95% confidence interval are shown overall and according to subgroup for association between the *MUC5B* promoter polymorphism variant allele and EHR-ARDS risk among participants in the BioVU cohort. All analyses were adjusted for age, sex, race (white versus non-white), presence of ARDS risk factors (exclusive of a risk factor when it is the subgroup definition), and presence of comorbid cardiac disorders (heart failure, acute myocardial infarction, or other coronary artery disease), but did not include an age × genotype interaction term.

### Impaired Oxygenation among Mechanically Ventilated BioVU Patients with *MUC5B* Promoter Polymorphism

Among BioVU participants who received invasive mechanical ventilation (regardless of EHR-ARDS status), rs35705950 variant carriers had more severe oxygenation impairment, with a mean -8 mmHg (95% CI: -15 to -1 mmHg) lower PaO_2_/FiO_2_ ratio per T-allele (p-value: 0.034, **Figure 4**). We observed a similar direction of effect among BioVU participants with EHR-ARDS (mean -6 mmHg per T-allele, 95% CI: -14 to +2, p-value: 0.14, **Figure 4**) which corresponded to increased odds for being in a more severe ARDS category by the Berlin Definition (OR: 1.23, 95% CI: 0.94 to 1.63, p-value: 0.14), although these associations did not meet our pre-specified level of statistical significance. We did not observe any such association between rs35705950 and oxygenation impairment in the VALID cohort (**e-Figure 6**).

**Figure 4:**
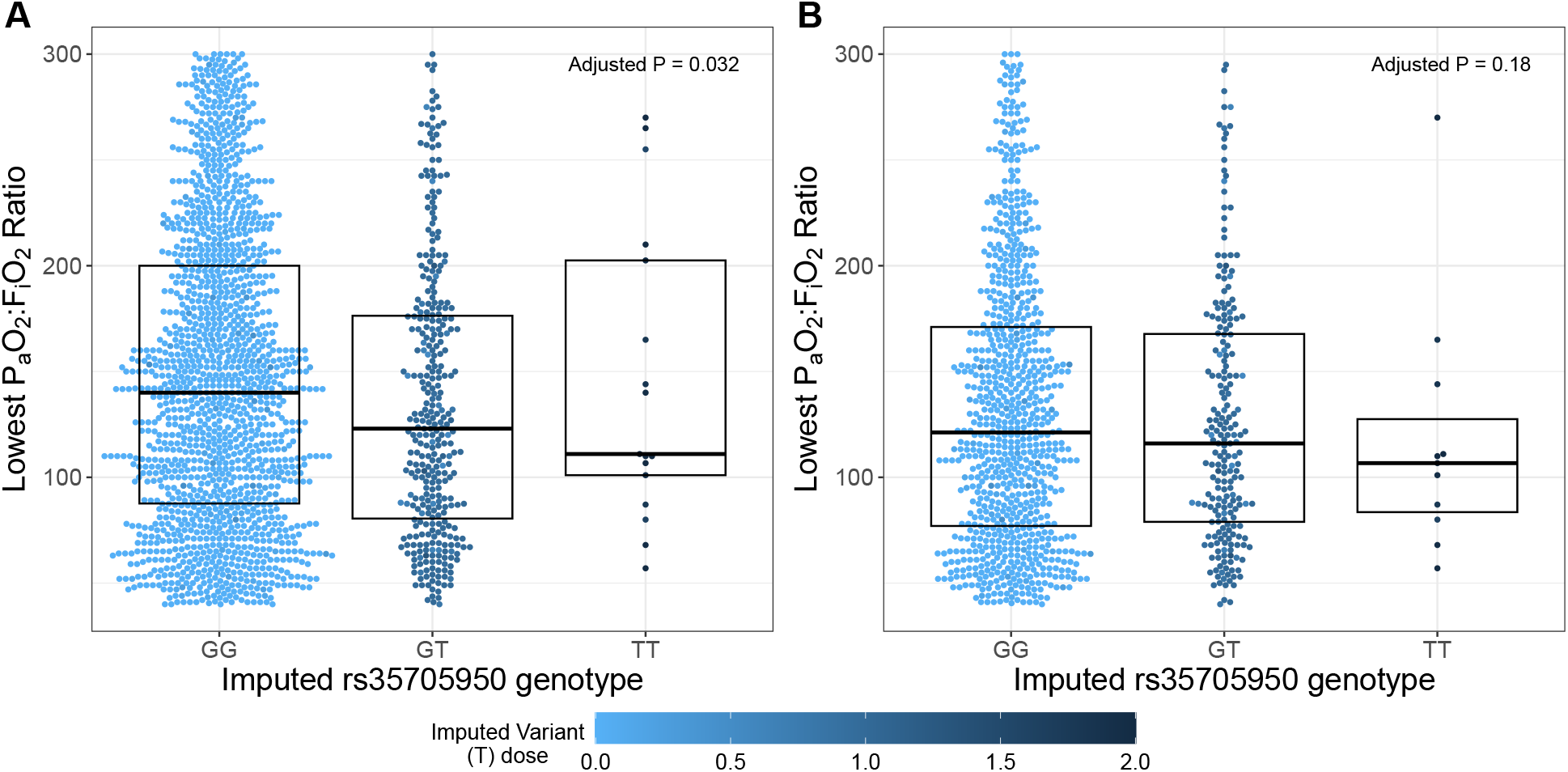
*MUC5B* promoter polymorphism is associated with more severe oxygenation impairment in mechanically ventilated patients and ARDS patients in the BioVU Cohort. Oxygenation impairment as measured by lowest P_a_O_2_:F_i_O_2_ ratio during the first seven days of hospitalization according to *MUC5B* promoter polymorphism (rs35705950) genotype among (**A**) all BioVU participants who received mechanical ventilation (N = 1,866) and (**B**) BioVU participants who met EHR-ARDS classifier (N = 1,056). Boxes indicate the median and interquartile range across each genotype. Colors for each dot indicate the imputed T-allele dosage, a continuous value ranging from light blue (T-allele dosage = 0.0; GG genotype) to dark blue (T-allele dosage = 2.0; TT genotype). Imputed genotypes were assigned based on T-allele dosage ranges of 0.0 to 0.5 (GG genotype), 0.5 to 1.5 (GT genotype), or 1.5 to 2.0 (TT genotype).

## DISCUSSION

### Summary of Findings

In this proof-of-concept study, we tested the utility of our de-identified longitudinal EHR to recapitulate a previously-observed genetic association with the *MUC5B* promoter polymorphism rs35705950 and ARDS risk in hospitalized adults. While our prior analysis in VALID reported an association between rs35705950 and ARDS among patients aged ≥50 years, our current study extends these findings in the VALID cohort by demonstrating a significant age × genotype interaction effect in nearly twice as many patients as the previous study cohort.^12^ These findings were additionally robust to adjustment for the presence of several ARDS risk factors and potentially confounding cardiac diagnoses. Complementary to our findings in VALID, in our de-identified BioVU cohort we found an association between rs35705950 and EHR-ARDS risk, although this association was generally consistent across all age ranges. We also observed a novel genotype-specific association with oxygenation impairment in BioVU. Each T-allele was associated with a mean 8 mmHg lower P_a_O_2_:F_i_O_2_ ratio among BioVU participants receiving invasive mechanical ventilation, and there was a directionally similar effect among those meeting our EHR-ARDS definition. Finally, our EHR-ARDS algorithm had similar levels of agreement with investigator-adjudicated ARDS as clinician documentation^9^ or inter-observer agreement between multiple clinicians,^23^ supporting the clinical validity of this EHR phenotype. The performance of our algorithm differed between cohorts, with a higher specificity and PPV in BioVU compared to VALID; this may reflect differences between cohorts in baseline demographics, severity of illness, and rates of ARDS risk factor diagnoses.

### Relationship to prior literature

The gel-forming mucin *MUC5B* is highly expressed in bronchial epithelium and has important roles in mucociliary clearance and control of bacterial infection and inflammation.^24,25^ Originally identified as a risk factor for interstitial lung disease,^26,27^ the promoter polymorphism rs35705950 lies 3 kilobases upstream of the transcription start site and is associated with transcriptional over-expression of *MUC5B* mRNA in distal airways and respiratory bronchioles.^26,28^ Potential mechanisms for the association between rs35705950 and ARDS may include increased risk for ARDS among patients with preclinical or undiagnosed interstitial lung disease, or *MUC5B* over-expression leading to mucociliary dysfunction and increased lung inflammation during acute illness.^29,30^ This study is the second to report an association between rs35705950 and ARDS risk in a general population of critically ill adults, although there is some overlap as both studies included patients from the VALID cohort. Our findings contrast those of a multinational genome-wide meta-analysis which reported a lower risk for hospitalization for rs35705950 T-allele carriers in patients with COVID-19,^31^ although potential explanations may include virus-specific effects of airway mucins, selection bias due to differences in self-isolation behaviors during the COVID-19 pandemic, or survivor bias among variant carriers who never developed interstitial lung disease.^32^

Prior efforts to identify ARDS from the EHR often focused on the role(s) of specific EHR data elements such as clinician documentation,^9,33^ chest radiograph report text,^8,34–37^ or radiographic images,^10^ with a focus on clinical applications such as implementing evidenced-based ARDS care or clinical trial recruitment. In contrast, our study integrated multiple data sources available in a widely-used EHR common data model including diagnosis codes, laboratory studies, and free-text chest radiograph reports.^15,38^ Sathe et. al. evaluated the performance of a conceptually similar EHR-ARDS classifier among adults hospitalized with COVID-19, and reported high agreement between their EHR-ARDS classifier and clinician-adjudicated ARDS (κ=0.85) in 175 manually reviewed patients.^11^ Li et. al. evaluated an EHR-ARDS classifier using laboratory values and chest radiograph report text across seven medical centers in a large multi-state health system, also reporting very high performance of their algorithm (sensitivity and specificity both above 90%) against manual adjudication for 150 patients.^39^ Out study had over 10-fold more patients with manually-adjudicated ARDS status than either previous study, therefore our results may provide more precise estimates of the real-world performance for a rules-based EHR-ARDS classifier.

### Strengths

Our VALID cohort is well-established in the critical care literature and provided nearly 2800 patients with extensive investigator-adjudicated ARDS phenotyping data.^9,12,13,40^ Similarly, BioVU is one of the largest institutional EHR biobanks in the United States and has yielded important insights into genetic drivers underlying a range of medical conditions.^*7,41,42*^ Both cohorts captured a broad swath of ARDS including patients with medical, surgical, and trauma admissions as well as those with both direct and indirect causes of lung injury. Additionally, rather than *a priori* excluding patients with heart failure, our EHR-ARDS classifier algorithm specifically allowed for patients with concomitant cardiogenic and noncardiogenic causes of pulmonary edema since cardiac comorbidities and volume overload are common in adults with

ARDS.^*8,43,44*^ Whereas many prior ARDS genetics studies were limited only to ICU patients,^4– 6,12,45,46^ we specifically included non-ICU patients in BioVU to mitigate selection bias as many patients with ARDS risk factors who ultimately do not develop the syndrome are managed outside of the ICU. The stronger association between the *MUC5B* promoter polymorphism and EHR-ARDS among the subgroup of mechanically ventilated BioVU participants suggests that our overall findings were not driven by non-critically ill participants. Although we used imputed *MUC5B* promoter genotypes, we observed very high correlation between genotypes determined by imputation and genotypes determined by polymerase chain reaction which were available from the prior study in VALID,^*12*^ so the likelihood of genotype misclassification in the BioVU cohort is low.

### Limitations

This study has some limitations. Although our use of two cohorts supports the internal validity of our findings, both the VALID and BioVU cohorts came from a single center. Therefore, the generalizability of our findings to patient populations outside our center remains to be assessed. Overlap between the cohorts is possible as institutional protocols to prevent re-identification of BioVU participants precluded us from specifically excluding VALID patients from the BioVU cohort. Over this study’s time frame our center admitted over 105,000 adults to these ICUs, most of whom (>85%) have never submitted DNA samples to BioVU. So although there may be some patients enrolled in both cohorts, the amount of overlap is likely low. We excluded patients with pre-existing ILD based on presence of relevant diagnosis codes prior to hospital discharge. While manual review of chest imaging might be more sensitive, particularly for preclinical ILD,^27^ the large numbers and de-identified nature of BioVU made manual review impractical. We used a directed acyclic graph to inform model design and identify potential confounding variables, however as with all observational cohort studies these analyses may be subject to residual confounding. Finally, our EHR-ARDS classifier used a simple rules-based algorithm which demonstrated acceptable sensitivity but only moderate PPV compared to investigator-adjudicated ARDS. Additional refinements to the EHR-ARDS classifier incorporating additional data elements, more intensive natural language processing of chest radiograph reports, or machine learning algorithms may improve performance in future studies.^47^

## CONCLUSION

The *MUC5B* promoter polymorphism was associated with an increased risk for ARDS among two parallel cohorts of at-risk hospitalized adults enrolled at a single academic medical center. Although age-related effect modification was only observed in the prospective VALID cohort, the de-identified BioVU biobank cohort identified a more consistent association between this polymorphism and EHR-ARDS risk across all ages, and a novel association with more impaired oxygenation among mechanically ventilated participants. Our study illustrates the complementary roles of these differing cohort enrollment strategies and highlights the potential for population-based EHR biobanks to enable future investigation into the genetic determinants of critical illness at large scales.

## Supporting information

Manuscript Supplement

## Data Availability

All data produced in the present study are available upon reasonable request to the authors.

## ABBREVIATIONS

ARDS: (Acute Respiratory Distress Syndrome)
EHR: (Electronic Health Record)
MUC5B: (Mucin 5B)
NPV: (negative predictive value)
PPV: (positive predictive value)
ILD: (Interstitial lung disease)
VALID: (Validating Acute Lung Injury biomarkers for Diagnosis)

