## Supplementary material for "Electronic health record biobank cohort recapitulates an association between the *MUC5B* promoter polymorphism and ARDS in critically ill adults": Manuscript Supplement

Online Supplementary Appendix

### SUPPLEMENTARY METHODS

#### A. Study Setting

Study participants came from Vanderbilt University Medical Center (VUMC), a private nonprofit academic medical institution based in Nashville, Tennessee, USA. VUMC has maintained an EHR since the mid-1990's and largely eliminated paper records in clinical care since 2004.[1,2] As of 2024, VUMC had 1,741 licensed beds across 7 hospitals, and annually manages over 3.0 million ambulatory visits, 204,000 emergency department visits, and 78,000 hospital discharges.

#### B. Vanderbilt Synthetic Derivative (SD)

The Synthetic Derivative is a de-identified database of clinical and related data derived from the VUMC's clinical systems that has been restructured for research.[1] Data is repurposed from VUMC's enterprise data warehouse and transformed to the SD using a set of custom Extract, Transform, and Load (ETL) pipelines to map data to the Observational Medical Outcomes Partnership (OMOP) common data model.[3] The de-identification of SD records was achieved primarily through the application of a commercial electronic program. The SD encompasses records for over 3.5 million unique individuals who have received care at VUMC dating back to the mid-1990s. Data types available in the SD include encounter and visit data, reimbursement codes, clinical notes and documentation, nursing records, medication records, and clinical laboratory test results. Additional details on the design and implementation of the SD have been published previously.[1]

#### C. BioVU DNA Biobank Repository

BioVU is Vanderbilt's de-identified biobank using DNA samples obtained from blood collected for clinical care that would have otherwise been scheduled to be discard after completion of clinical testing. BioVU uses a de-identification program linking participants' DNA samples to their de-identified EHR data in the SD using a unique research identifier code generated for each participant via a Secure Hash Algorithm. A full description of the BioVU program eligibility, consent methods, DNA extraction methods, genotyping, and ethical protections have been published previously.[4–8]

#### D. Investigator Access to Study Data

The primary author (VEK) had full access to all patient and clinical data available for both the VALID cohort and in the Synthetic Derivative data warehouse. QF and WQW provided access to genetic data for the BioVU cohort, and LBW provided access to genetic data for the VALID cohort.

#### E. Study Censoring Date

We censored all data in the Synthetic Derivative data warehouse entered either before October 1, 2000 or after July 15, 2019 (in the randomly re-assigned SD date space). The electronic health record at Vanderbilt dates back to the early 1990s,[2] but in practice the overwhelming majority of data (98.9%) in our cohort were for diagnosis and laboratory tests occurring after January 2000.

### F. Quality Control of Study Data

**Patient-level quality control.** We excluded patients with missing data for date of birth, recorded sex, or those that had zero visit or diagnosis code records present in the EHR after January 1, 2000. We also excluded patients denoted as being “dummy” or “test” records in the EHR. We included patients who had race or ethnicity coded as “Unknown / Not reported” as some patients choose not to provide a self-identified race or ethnicity when being registered for a care visit.

**ICD code data quality control.** The Synthetic Derivative is refreshed semi-annually, therefore some data are subject to change as they are finalized, amended, corrected, or updated in the medical record. Pertinent to this study, ICD diagnosis codes are typically entered into the enterprise data warehouse by practitioners, but can be changed during finalization by coding specialists several days or weeks later. Our experience and that of the VICTR Big Data staff has been that ICD codes and problem lists have relatively low volatility over time.[1]

**Laboratory value data quality control.** Clinical laboratory test records in the Synthetic Derivative data warehouse are imported from VUMC’s laboratory information system (Oracle Cerner, North Kansas City, MO). Similar to our experience with diagnosis codes, we have found relatively low volatility for these results once they are finalized in the EHR.[1]

**Ascertainment of Death Status.** Death records are entered into the VUMC Synthetic Derivative data warehouse from multiple potential sources, including (i) inpatient deaths occurring at our center; (ii) deaths occurring outside our center reported to our Medical Records Operations service by a decedents’ family members, outside facilities, administrative staff, or clinicians; (iii) deaths reported to institutional disease-related registries, or (iv) from the Social Security Death Index (SSDI) which was available through December 31, 2016. When date of death was unavailable in the Synthetic Derivative (only for death records from the SSDI), we used date of last contact as measured by the date of last diagnosis code, visit, or clinical communication event in our EHR. Patient who were not known to be deceased based on the Synthetic Derivative were assumed to be alive at the date of study censoring (July 15, 2019).

### G. *MUC5B* Promoter Genotyping

**Quality Control.** Pre-imputation genotyping quality control measures included filtering single-nucleotide polymorphisms (SNP) with <98% call rate or <1.0% minor allele frequency, discordance between reported sex and X-chromosome zygosity, pairs with cryptic relatedness by $\hat{\pi} > 0.2$, or extreme deviation from Hardy-Weinberg equilibrium with $p < 1.0\times10^{-6}$ in ARDS controls and $p < 1.0\times10^{-10}$ in ARDS cases.

***MUC5B* Promoter Polymorphism Quality Control**. The observed imputation certainty ($R^2$) in the overall BioVU cohort (N = 94,864) was 0.9011. The observed imputation $R^2$ in the VALID cohort (N = 2,884) was 0.9935. In both cohorts this was well above generally accepted quality control measures for use of imputed variants in genetic association studies.[9,10]

**SUPPLEMENTARY FIGURES**

**A. e-Figure 1. Causal Diagram for Primary Analysis**

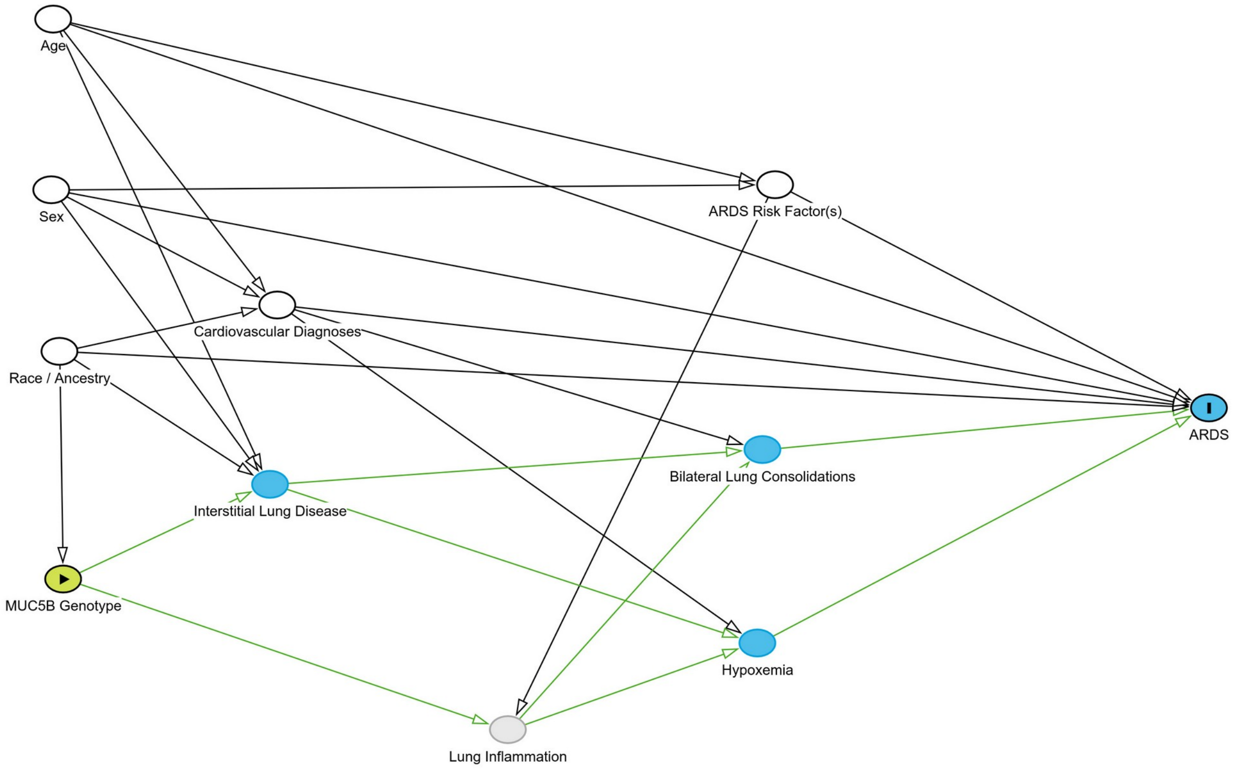

Directed acyclic graph illustrating causal relationships between exposure, outcome, and other variables used in analyses. Yellow circle with the right-pointing triangle represents exposure of interest (MUC5B Genotype). Blue circle with "I" indicates outcome of interest (ARDS). White circles represent potential confounding variables which were adjusted in the primary analysis (Age, Sex, Race, ARDS Risk Factors, and Cardiovascular Diagnoses). Grey square represents variables which were used for selection / exclusion (Interstitial lung disease). Grey circle represents unobserved variables which were not adjusted for in the analyses. (Lung Inflammation). Remaining blue circles represent other observed ancestors of the outcome which were not adjusted for in the primary analysis (Bilateral Lung Consolidations, Hypoxemia). Green lines represent the causal path between the exposure of interest (MUC5B) and outcome (ARDS). Created using R package *dagitty* version 3.1.[11] Available online at: https://dagitty.net/mW3pBhUsC

### B. e-Figure 2. Flow Diagrams for Study Cohorts

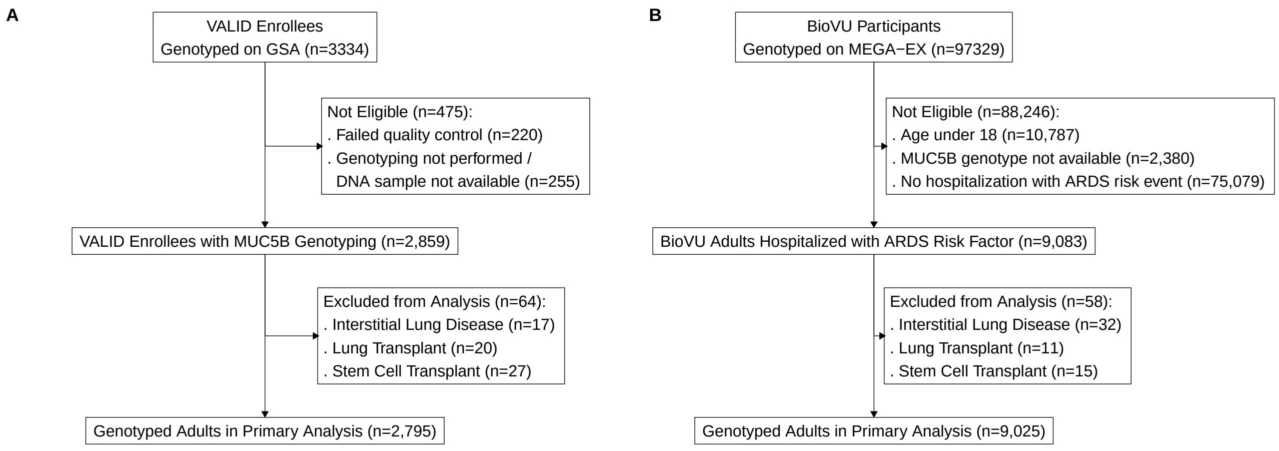

Flow diagrams for (A) VALID prospective biomarker cohort and (B) BioVU de-identified DNA biobank cohort. GSA: Illumina® Global Screening Array. MEGA-EX: Illumina® Expanded Multi-Ethnic Global Array. VALID: Validating Acute Lung Injury biomarkers for Diagnosis cohort study.

**C. e-Figure 3. Distribution of Imputed *MUC5B* Promoter Polymorphism in Study Populations**

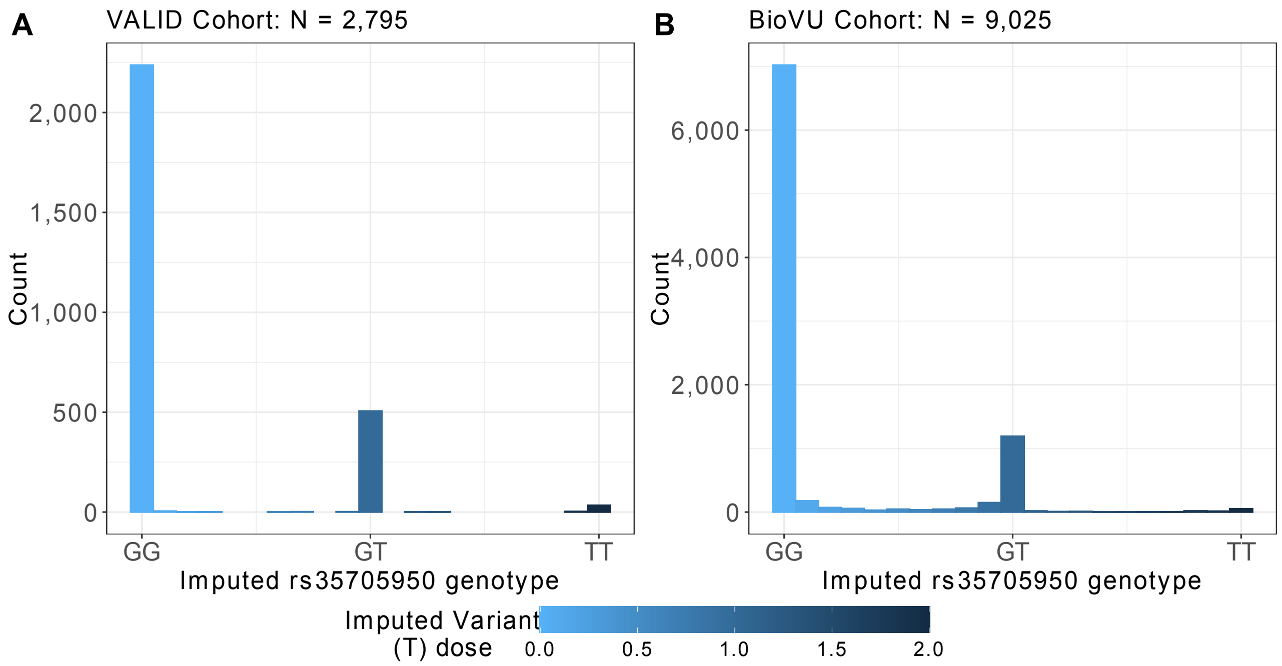

Histograms of imputed *MUC5B* promoter polymorphism (rs35705950) variant (T) allele dosages among (A) VALID patients and (B) BioVU participants. VALID: Validating Acute Lung Injury biomarkers for Diagnosis.

### D. e-Figure 4. In-Hospital Survival for ARDS and EHR-ARDS based on Berlin Severity

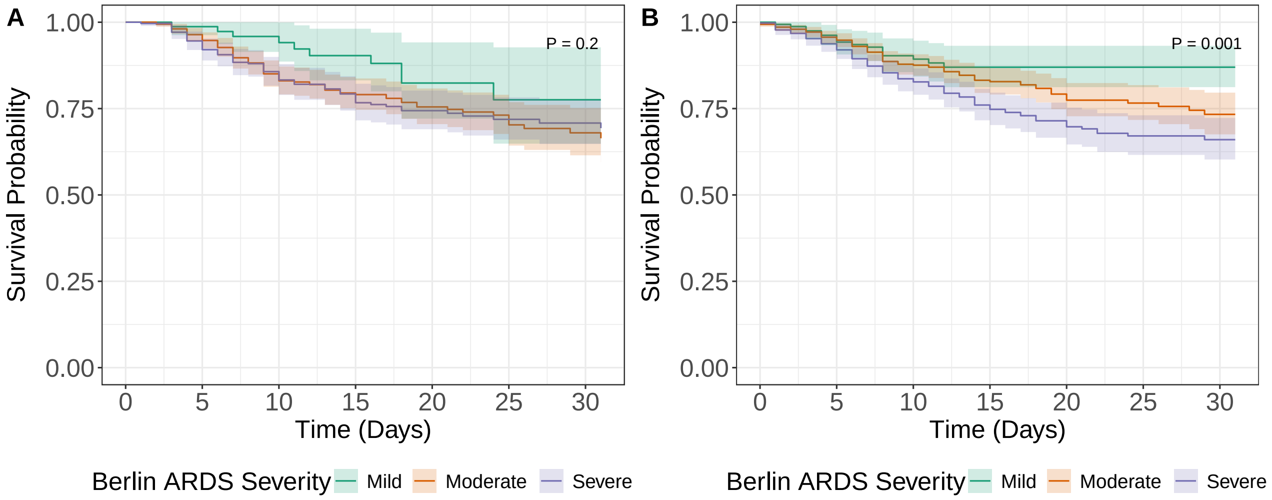

**A** — At Risk (Events)

| | 0 | 5 | 10 | 15 | 20 | 25 | 30 |
|---|---|---|---|---|---|---|---|
| Mild | 78 (0) | 73 (1) | 54 (4) | 41 (6) | 25 (9) | 16 (10) | 15 (10) |
| Moderate | 363 (0) | 342 (19) | 254 (57) | 174 (67) | 111 (73) | 78 (79) | 50 (81) |
| Severe | 277 (0) | 262 (22) | 214 (45) | 156 (59) | 108 (63) | 70 (66) | 54 (67) |

**B** — At Risk (Events)

| | 0 | 5 | 10 | 15 | 20 | 25 | 30 |
|---|---|---|---|---|---|---|---|
| Mild | 160 (0) | 146 (9) | 91 (15) | 49 (17) | 29 (17) | 22 (17) | 11 (17) |
| Moderate | 494 (3) | 441 (25) | 318 (55) | 211 (69) | 135 (79) | 83 (80) | 56 (83) |
| Severe | 402 (1) | 372 (32) | 277 (66) | 183 (88) | 124 (98) | 82 (102) | 56 (103) |

Plots of Kaplan-Meier estimators for 30-day in-hospital survival based on ARDS severity among (**A**) VALID patients with Berlin ARDS by investigator-adjudication (N = 718), and (**B**) BioVU participants meeting the EHR-ARDS classifier definition (N = 1,056).[12] Patients/participants were group by Mild ARDS (green lines, lowest $P_aO_2$:$F_iO_2$ ratio between 201 and 300), Moderate ARDS (orange lines, lowest $P_aO_2$:$F_iO_2$ ratio between 101 and 200), and Severe ARDS (purple lines, lowest $P_aO_2$:$F_iO_2$ ratio 100 or less). P-values indicate significance across all three groups using the log-rank test. Tables underneath each plot indicate number of patients/participants At Risk and (Number of Deaths) at five-day intervals. VALID: Validating Acute Lung Injury biomarkers for Diagnosis.

**E. e-Figure 5. Subgroup Analysis of *MUC5B* Promoter Polymorphism and ARDS Risk in VALID Cohort**

| Subgroup | GG | GT | TT | OR (95%CI) |
|---|---|---|---|---|
| All Patients | 2,245 | 513 | 37 | 1.11 (0.90 to 1.37) |
| Age | | | | |
| Age >= 50 | 1,397 | 332 | 24 | 1.24 (0.95 to 1.62) |
| Age < 50 | 848 | 181 | 13 | 0.92 (0.65 to 1.31) |
| Sex | | | | |
| Males | 1,348 | 311 | 20 | 0.98 (0.74 to 1.30) |
| Females | 897 | 202 | 17 | 1.29 (0.92 to 1.79) |
| Clinical | | | | |
| Mechanical Ventilation | 1,692 | 406 | 30 | 1.02 (0.82 to 1.28) |
| Survived Hospitalization | 1,887 | 421 | 31 | 1.03 (0.80 to 1.32) |
| Inpatient Death | 358 | 92 | 6 | 1.45 (0.90 to 2.35) |
| ARDS Risk Factor | | | | |
| Aspiration | 295 | 63 | 5 | 0.94 (0.56 to 1.55) |
| Pancreatitis | 109 | 26 | 1 | 0.78 (0.25 to 2.39) |
| Pneumonia | 989 | 237 | 16 | 1.14 (0.87 to 1.51) |
| Sepsis | 1,055 | 236 | 21 | 1.05 (0.80 to 1.39) |
| Shock | 1,534 | 355 | 24 | 1.21 (0.95 to 1.54) |
| Trauma | 785 | 192 | 10 | 1.06 (0.76 to 1.46) |

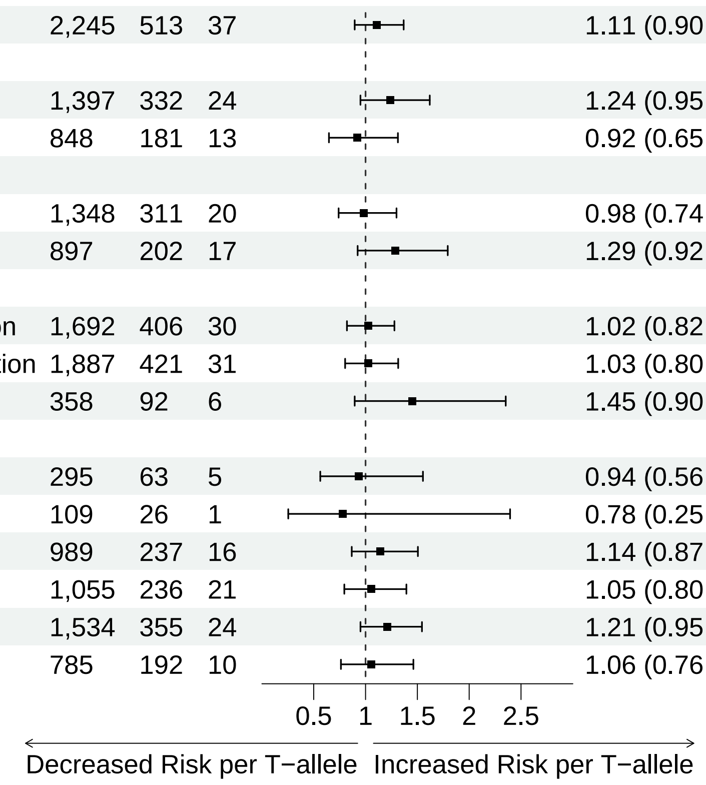

The odds ratio and 95% confidence interval are shown overall and according to subgroup for association between the *MUC5B* promoter polymorphism variant allele and ARDS risk among patients from the VALID cohort. All analyses were adjusted for age, sex, race (white versus non-white), presence of ARDS risk factors (exclusive of a risk factor when it is the subgroup definition), and presence of comorbid cardiac disorders (heart failure, acute myocardial infarction, or other coronary artery disease), but did not include an age × genotype interaction term. VALID: Validating Acute Lung Injury biomarkers for Diagnosis.

**F. e-Figure 6. *MUC5B* promoter polymorphism was not associated with oxygenation impairment among mechanically ventilated patients in the VALID cohort**

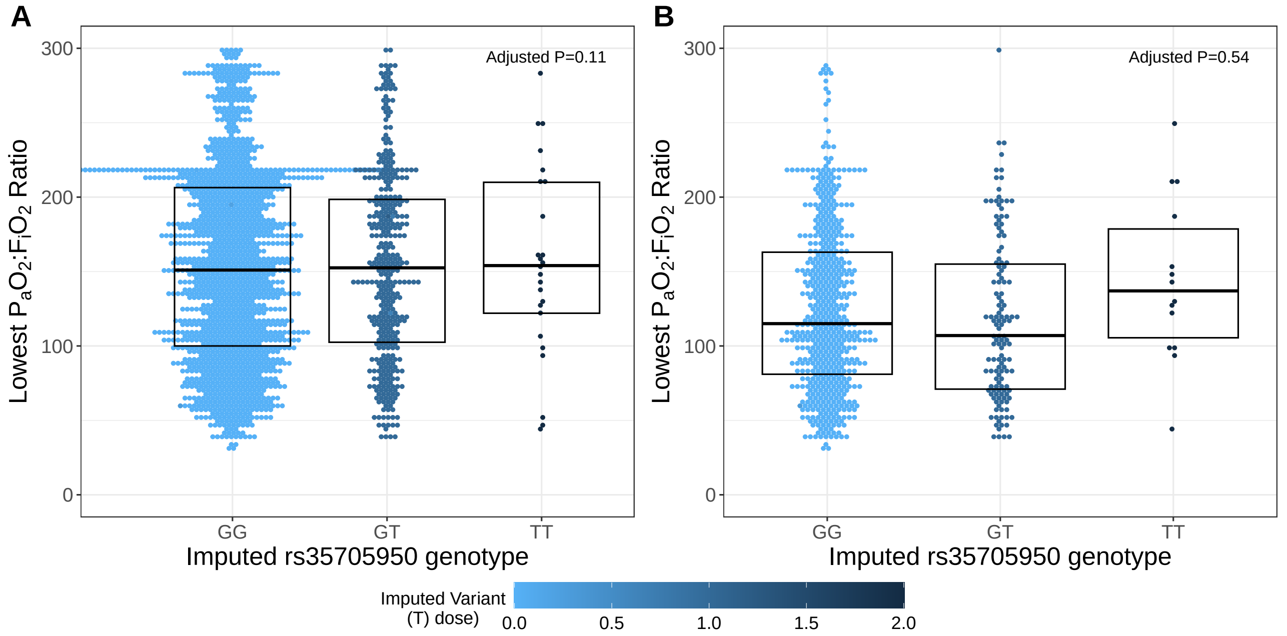

Oxygenation impairment as measured by lowest $P_aO_2$:$F_iO_2$ ratio during the first four intensive care unit days according to *MUC5B* promoter polymorphism (rs35705950) genotype among (**A**) all VALID patients who received mechanical ventilation (N = 2,128) and (**B**) VALID patients with investigator-adjudicated Berlin ARDS (N = 718). Boxes indicate the median and interquartile range across each genotype. Colors for each dot indicate the imputed T-allele dosage, a continuous value ranging from light blue (T-allele dosage = 0.0; GG genotype) to dark blue (T-allele dosage = 2.0; TT genotype). Imputed genotypes were assigned based on T-allele dosage ranges of 0.0 to 0.5 (GG genotype), 0.5 to 1.5 (GT genotype), or 1.5 to 2.0 (TT genotype). VALID: Validating Acute Lung Injury biomarkers for Diagnosis.
